# Digital Exclusion as a barrier to accessing healthcare: A summary composite indicator and online tool to explore and quantify local differences in levels of exclusion

**DOI:** 10.1101/2023.07.12.23292547

**Authors:** Paul Mee, Mark Gussy, Phil Huntley, Amanda Kenny, Theo Jarratt, Nigel Kenward, Derek Ward, Aiden Vaughan

**Affiliations:** Lincoln Institute for Rural and Coastal Health, College of Health and Science, University of Lincoln, UK; School for Data Science and Computational Thinking, Stellenbosch University, South Africa; La Trobe Rural Health School, College of Science, Health and Engineering, La Trobe University, Australia; Public Health Division, Lincolnshire County Council, Lincoln, UK; NHS Lincolnshire Integrated Care Board, Lincoln, UK; United Lincolnshire Hospitals NHS Trust, Lincoln, UK

**Keywords:** Digital Exclusion, Access, Composite indicator, Marginalisation, Inequity, Social Deprivation

## Abstract

Existing disparities in digital access were exacerbated with the accelerated shift to online provision of services during the COVID-19 pandemic, particularly for already disadvantaged groups. Metrics to quantify relative local differences in levels of digital exclusion are a necessary pre-requisite for the targeting of interventions to address these disparities. In this study we developed a composite indicator and an interactive dashboard ‘The Lincolnshire Digital Health Toolkit’ to explore digital exclusion in Lincolnshire, UK. To develop the indicator, individual variables were normalised and aggregated, intra-variable correlations explored, and factor analysis used to determine variable weightings. Three underlying factors were identified that explained a significant proportion of the variance in the original variables, the first two predominantly related to socioeconomic deprivation and lack of activity. In general, coastal areas in the east of the county had higher levels of digital exclusion, with significant local variation particularly within urban areas. Long travel times to reach medical facilities are an additional barrier in some communities. The toolkit has been used to support the evidence-based geographic targeting of interventions to address barriers to accessing digitally based health information and services. The impact of digital exclusion must be addressed to reduce marginalisation and isolation. The Lincolnshire Digital Health Toolkit provides a novel composite metric tailored to the conditions of this largely rural county and an interactive dashboard to support decisions on resource allocation.

## 1. Introduction

The provision of key services online and the impact of inequity in digital access is attracting increasing international interest. The COVID-19 pandemic and lockdown measures resulted in a step-change in the provision and adoption of digital tools. In the United Kingdom (UK) and other high income countries online provision became the norm for medical appointments, engagement with social support services, and education at all levels. As a consequence, pre-existing inequalities in access and take-up of digital technology were exacerbated [1]. While the pandemic was a once in a lifetime event, globally there has been a major change in the adoption of digital technology with evidence pointing towards sustained changed patterns of remote or flexible working. Given these important trends, there is a critical need to explore the significant section of the population who face digital exclusion. These groups already faced major digital disadvantages prior to the pandemic, during the pandemic and in a post-pandemic world they have experienced an increased level of marginalisation that amplified their exclusion [2–4].

Ofcom, the UK communications regulator, describe digital exclusion as having three inter-linked dimensions relating to access, ability and affordability [5]. Access describes exclusion due to a lack of internet provision at home or elsewhere. Affordability is closely linked and relates to the financial cost of accessing the internet or purchasing appropriate devices for access. Ability describes barriers due to lack of digital literacy. Digital exclusion results in major disadvantage, marginalisation, and inequality, particularly for those who are geographically isolated and have health-related limitations or impairment [3]. There is a lack of tools to determine the relative digital exclusion of people at fine-grained local geographic levels which makes it challenging to develop targeted interventions to address this.

Those living in geographically isolated communities may experience inequitable access to services, often due to a lack of public transport. Geographic barriers are further intensified for individuals with a disability or illness which restricts their mobility, for whom face to face access to a medical practice can be challenging [6]. Online service provision is often put forward as a solution for those facing geographic isolation, but lack of digital connectivity, low levels of digital literacy, and a lack of access to digital resources limits the take-up of such services for these groups [7]. UK data indicates that only 78% of individuals with a health-related limitation or impairment make use of the internet compared to 94% of individuals in the general population [7]. While there is a lack of granular data on internet usage amongst people who face both ill-health or impairment and geographic isolation, it could be hypothesised that the internet access differences would be even wider.

Digital inclusion can be characterised as a basic human right [8] with those who are digitally excluded facing major challenges including in seeking employment, accessing banking and managing money, and taking-up online learning opportunities [9]. There is a clear connection between digital exclusion and social isolation [10–12]. A recent report defines the concept of a Minimum Digital Living Standard (MDLS) as *“having accessible internet, adequate equipment, and the skills, knowledge and support people need. It is about being able to communicate, connect, and engage with opportunities safely and with confidence”* [13].

Interventions which promote digital inclusion need to be tailored to the needs of the individual or community and the type of services they need to access. In this paper, we describe the development of a composite indicator and associated online tool to quantify the relative digital exclusion of each Lower-layer Super Output Area (LSOA) in the County of Lincolnshire in the UK. Lincolnshire is comprised of urban, rural and coastal communities with almost a quarter (23%) of the population aged over 65 compared to 18% across the whole of England [14]. The demography and overall low population density of the county leads to particular challenges for older people in accessing health and social care and public transport. This study addresses a methodological gap in the development of composite indicators which quantify digital exclusion. We describe the approach taken to choose the underlying indicator variables and statistical methods used to normalise and aggregate them into a single composite index balancing the contribution of the underlying constructs within the individual indicators. The composite indicator and its method of development has potential applicability across the UK and internationally. The Lincolnshire Digital Health Toolkit (https://lhih.org.uk/lincolnshire-digital-health-toolkit/) provides a web-based user interface to explore geographic variations in digital exclusion.

## 2. Background

Lincolnshire is a large rural and coastal, geographically dispersed county, which poses challenges for health and other service provision [14]. The UK Chief Medical Officer highlighted the lack of digital infrastructure as a key challenge in coastal communities in his 2021 annual report, and included Lincolnshire as a case study [15]. Lincolnshire has an ageing population in a widely rural area with the more deprived areas of Lincolnshire seen on the coast and the inner towns and cities.

Lincolnshire County Council (LCC) worked with other Local authorities on the ‘Digital Access for All’ study [16]. As a Council, LCC were committed to addressing digital exclusion for adults with social care needs. A framework was developed showing the steps which local authorities and health and social care providers might take to develop locally tailored digital inclusion strategies. Key to the implementation of such a framework is the development of metrics that quantify levels of exclusion and identify individuals and local geographic areas of greatest need. A variety of underlying factors have been identified which contribute to digital exclusion in addition to limitations in the provision of physical access to online resources. These include age, gender, income level, type of or lack of employment and level of educational attainment [17].

### 2.1 Composite Indicators

Many composite indicators have been developed across a multitude of domains including deprivation, national economic performance, human development, and health service delivery [18–20]. These indicators are potentially useful in analysing the immediate and longer-term impact of policy change. Examples of these include: the Social Wellbeing index [22, 23], which seeks to present a nuanced measure of an individual’s quality of life encompassing the strength of their social connections, the UK indices of multiple deprivation includes factors related to material deprivation in addition to measures of health, education status and crime [24] and the United Nations Multidimensional Poverty Index, which provides an international comparison of multiple deprivations associated with health, education and standard of living [25]. However, poorly constructed indices may lead to misleading policy messages and in turn ineffective policy development and implementation[21]

Criticism has been directed at a lack of methodological transparency in the development of composite indices which can lead to users being unable to properly understand the rationale behind their development [20]. Significant issues include the rationale for selecting the component variables and the methods used to apply relative weightings to them in the derivation of the composite. If a composite index includes several indicators which describe the same underlying construct, and are thus highly correlated, there is a risk that these constructs may dominate other components in the overall score [20].

The Digital Exclusion Risk Index (DERI) Tool [26] was developed by the Greater Manchester Combined Authority research team in the UK to enable local authorities to better understand digital exclusion at a local level in order to target interventions to the areas of most need. The DERI tool combines a number of indicators to provide an overall DERI score for each Lower-layer Super Output Area (LSOA)^1^ across England, with visualisations of the relative scores provided for all LSOA’s in Greater Manchester. The overall score is a composite measure of the risk of digital exclusion ranging from 0 (low risk) to 10 (high risk) based on nine individual indicators^2^. Underlying indicators are grouped into three broad categories describing demography (age structure), deprivation, and broadband access. Indicators are standardised on a range of 0 to 10, they are then combined in to three composites: age, broadband and deprivation using an empirical weighting scheme. The three composites are then given equal weighting in the final composite [27].

The Lincolnshire Digital Health Toolkit has been created in a bid to reduce digital exclusion in Lincolnshire by highlighting areas at greatest risk of being left behind as a result of digitalisation by including indicators that give greater granularity and context to Lincolnshire and its population. The DERI tool does not directly take account of relative differences in the levels of digital literacy, that is the skills and confidence individuals have in accessing digital services. For this reason, Experian Mosaic indicators which take these factors into account were included in our new composite index. Experian [28] is a commercial organisation that produces small-area indicators of lifestyle, behaviour and socio-demographics, these are mainly used for market segmentation. The use of these Experian geo-demographic indicators for health research in the UK has been increasing in recent years. They can be used to identify health-related risk factors which can in turn be used to target limited resources to areas of greatest need [29]. Examples of their application include: predicting smoking prevalence [30] and the likely effectiveness of smoking cessation interventions [31], to assess the completeness and representativeness of lung cancer data [32] and to understand the associations between multimorbidity and sociodemographic characteristics [33]. The indicators are derived using a proprietary modelling strategy in which data from surveys of representative samples of the population are extrapolated to the whole population based on local demographic characteristics.

## 3. Methods

The indicator variables representing different dimensions of digital exclusion (Table 1) were collated for each LSOA in Lincolnshire. The choice of variables was based on a review of the literature and a series of consultations with stakeholders with knowledge of the sociodemographic and other factors acting as barriers to accessing services for the local population.

**Table 1:** Individual Indicators used to create the composite indicator for digital exclusion.

| Code | Description |
| --- | --- |
| DER | The percentage of the LSOA's population who are considered either D or E on the NRS<sup>3</sup> social grade. |
| GPC | Recipients of Guaranteed Pension Credit<sup>4</sup> (Rate per 1000 of LSOA population aged 65 and over). Pension Credit gives extra money to help with living costs if for those over the state pension age and on a low income. |
| INT | Average download speed in LSOA (Mb/sec)<sup>5</sup> |
| LAC | Percentage of population of the LSOA whose daily activity is severely limited<sup>6</sup> |
| NQR | The percentage of residents aged over 16 in the LSOA who have no qualifications<sup>6</sup> |
| UNP | The percentage of those aged over 16 in the LSOA who are unemployed and claiming benefits<sup>6</sup> |
| EXP | A composite of five Experian Mosaic indicators<sup>7</sup> showing the estimated level of competence in the use of email, smartphone, instant messaging, social networking or the internet amongst the LSOA population. |

Our step-wise approach to develop the composite indicators followed the strategy laid out by Mazziotta and Pareto [34]. In this approach it is recommended that each individual indicator should be positively correlated with the concept that is being represented in the overall composite. In our composite of ‘digital exclusion’ we want the LSOA ranked as 1 to be the most digitally excluded and the highest ranked LSOA to be the least digitally excluded. Thus, based on our causal hypotheses we postulate that for example an LSOA with a high percentage of individuals in NRS social class D or E will have a high level of digital exclusion (and a low ranking) hence we multiply the raw indicator by -1 to reflect that. By contrast a high value for the download speed indicator would be associated with a low level of exclusion and a high ranking in the composite hence we do not multiply this indicator by -1. After adjusting the polarity where necessary each variable was normalised with a mean value of 0 and a standard deviation of 1 to bring the indicators to the same standard, by transforming them into pure, dimensionless, numbers.

Two composites were developed using the methodology described below. The first, the ‘Digital Exclusion index’, included component variables representing factors associated with sociodemographic deprivation, local digital infrastructure and estimates of the level of competency in particular digital skills. The second was a composite of variables describing the average travel times by car or public transport to access a medical practice or pharmacy. Our rationale in developing these two indicators is that those with easy access to a medical facility will to some extent, have the means to overcome digital exclusion, whilst those in more remote areas will not have this advantage. The details of the variables included in the two composites and the codes used to describe them is shown in Tables 1 and 2.

**Table 2:** Individual Indicators used to create the composite indicator describing travel time to access a medical practice or pharmacy.

| Code | Description |
| --- | --- |
| C15 | Percentage of the population of the LSOA living within 15 minutes travel by car of a medical practice<sup>8</sup> |
| G15 | Percentage of the population of the LSOA living within 15 minutes travel by public transport of a medical practice<sup>8</sup> |
| G30 | Percentage of the population of the LSOA living within 30 minutes travel by public transport of a medical practice<sup>8</sup> |
| P15 | Percentage of the population of the LSOA living within 15 minutes by car of a pharmacy.<sup>8</sup> |

For the variables comprising the Experian (EXP) indicators we used the same methodology to initially create a composite from the component variables. We then included the resultant composite in the overall composite of digital exclusion.

### 3.1 Selection and derivation of Indicators

To investigate the degree of co-linearity between variables, correlation plots were derived showing the pairwise scatter plots overlaid with best fit regression lines and the values for the Pearson’s rank correlation coefficient (Figure 1) [36]. A correlation network diagram was developed again using the Pearson’s rank correlations showing which sub-groups of indicators were mutually correlated (Figure 2). Pearson’s correlation coefficient is a test statistic that measures the statistical relationship, or association, between two continuous variables. Coefficient values can range from +1 to -1, where +1 indicates a perfect positive relationship, -1 indicates a perfect negative relationship, and a 0 indicates no relationship exists.

**Figure 1:**
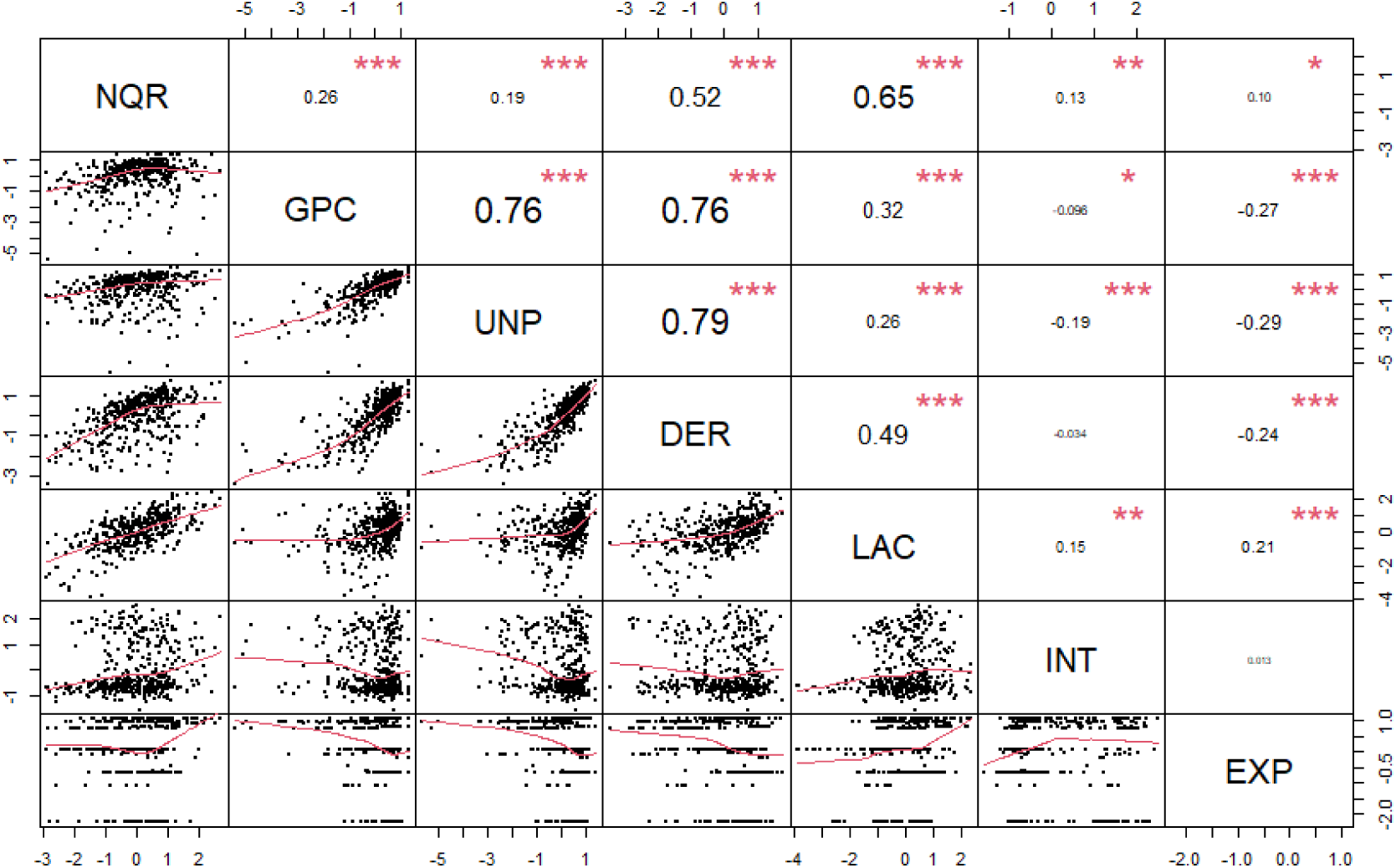
Summary correlation plot for the original indicator variables. Scatter plots of pairs of indicators are shown below the diagonal and the Pearson’s correlation coefficient (ρ) values are shown above the diagonal. p-values for the correlation are indicated as (* = <0.05, ** = <0.01, *** = <0.001). Indicator codes are described in Table 1.

**Figure 2:**
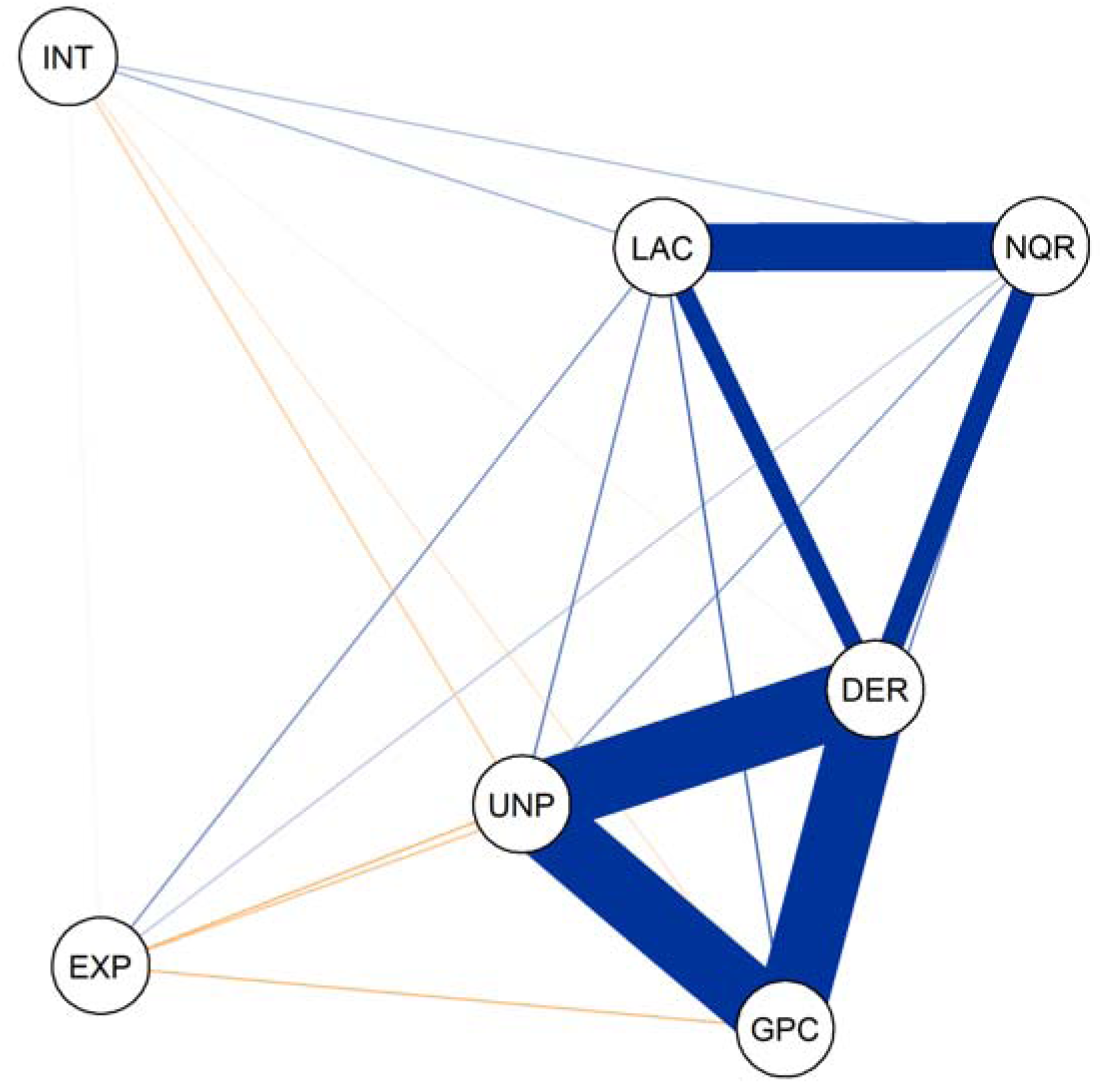
Correlation network diagram for the normalised indicator variables, the thickness of the lines linking pairs of nodes indicates the strength of the correlations pair of variables. Blue lines indicate a positive correlation and yellow lines a negative correlation between pairs of indicators. Indicator codes are described in Table 1.

Factor analysis (FA) is a statistical method akin to Principal Component Analysis (PCA) but operates under a unique model. While PCA focuses on linear combinations of data, FA assumes that observed data stem from underlying factors, with variance attributed to common and unique factors. The model expresses original variables in terms of a smaller set of uncorrelated common factors and unique factors. In the approach applied here, PCA is employed to extract the primary principal components, treating them as factors and disregarding the rest [37]. This simplifies analysis and is widely used in constructing composite indicators [21,38].

All analyses were carried out using the R statistical software package. [39] The correlation plot was generated using a customised version of the chart correlation function in the Performance Analytics package [40]. The correlation network diagram was generated using the qgraph package [41]. Weightings were obtained using the Compind package [42]. All maps were produced using the leaflet package [43]. All code is freely available for download from the following GitHub repository https://github.com/Paul-Mee/digex_linc.

## 4. Results

The correlation plot (Figure 1) and the correlation network diagram (Figure 2) describe the pairwise correlations between the eight indicator variables used to form the composite indicators. Three of these variables GPC (claiming of guaranteed pension credit), UNP (Unemployment), and DER (D or E socio-economic status) show a high degree of mutual correlation (correlation coefficients (ρ) GPC-UNP=0.76, GPC-DER=0.76, UNP-DER=0.79). The correlation between LAC (Low levels of activity) and NQR (lack of qualifications) is also high (ρ = 0.65) and these two are moderately correlated to DER (ρ LAC-DER=0.49, NQR-DER=0.52). These variables are not highly correlated with the other variables. EXP (composite of Experian indicators) and INT (average download speed) show a low degree of correlation with any of the other indicator variables.

The map of all LSOA’s (Figure 3) shows that the easternmost coastal areas of the county experience the highest degree of digital exclusion with a shift towards lower levels of exclusion towards the west.

**Figure 3:**
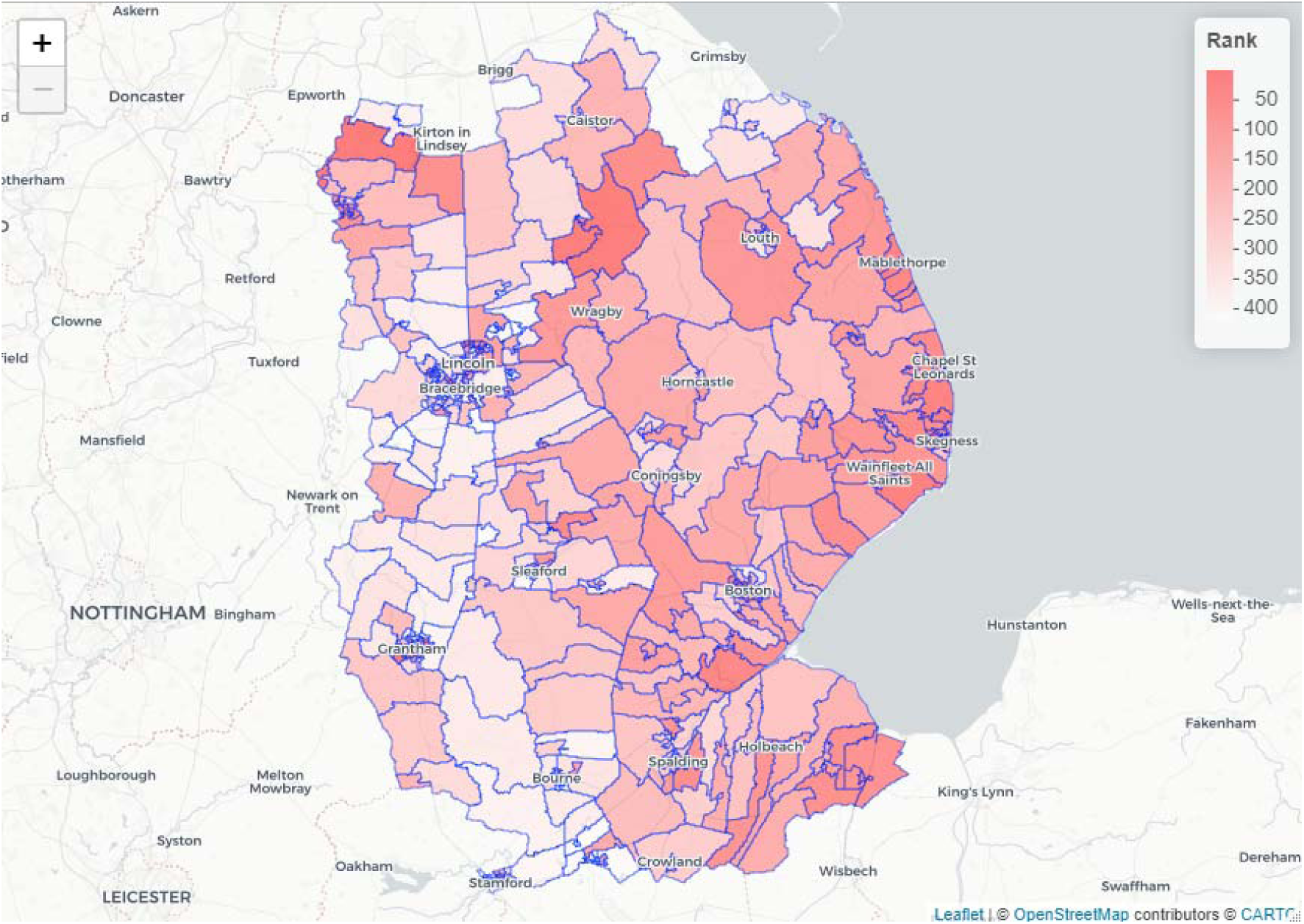
Map of Lincolnshire showing the relative rankings of the LSOA’s using factor analysis derived composite indicator scores for digital exclusion ranging from 1 (highest level of exclusion – dark red) to 420 (lowest exclusion – white).

In contrast the map of transport exclusion (Figure 4) shows that there are LSOAs throughout the county where individuals have relatively long travel times to access a medical facility or pharmacy. As would be expected the exception to this is for the LSOA’s situated in the main urban areas.

**Figure 4:**
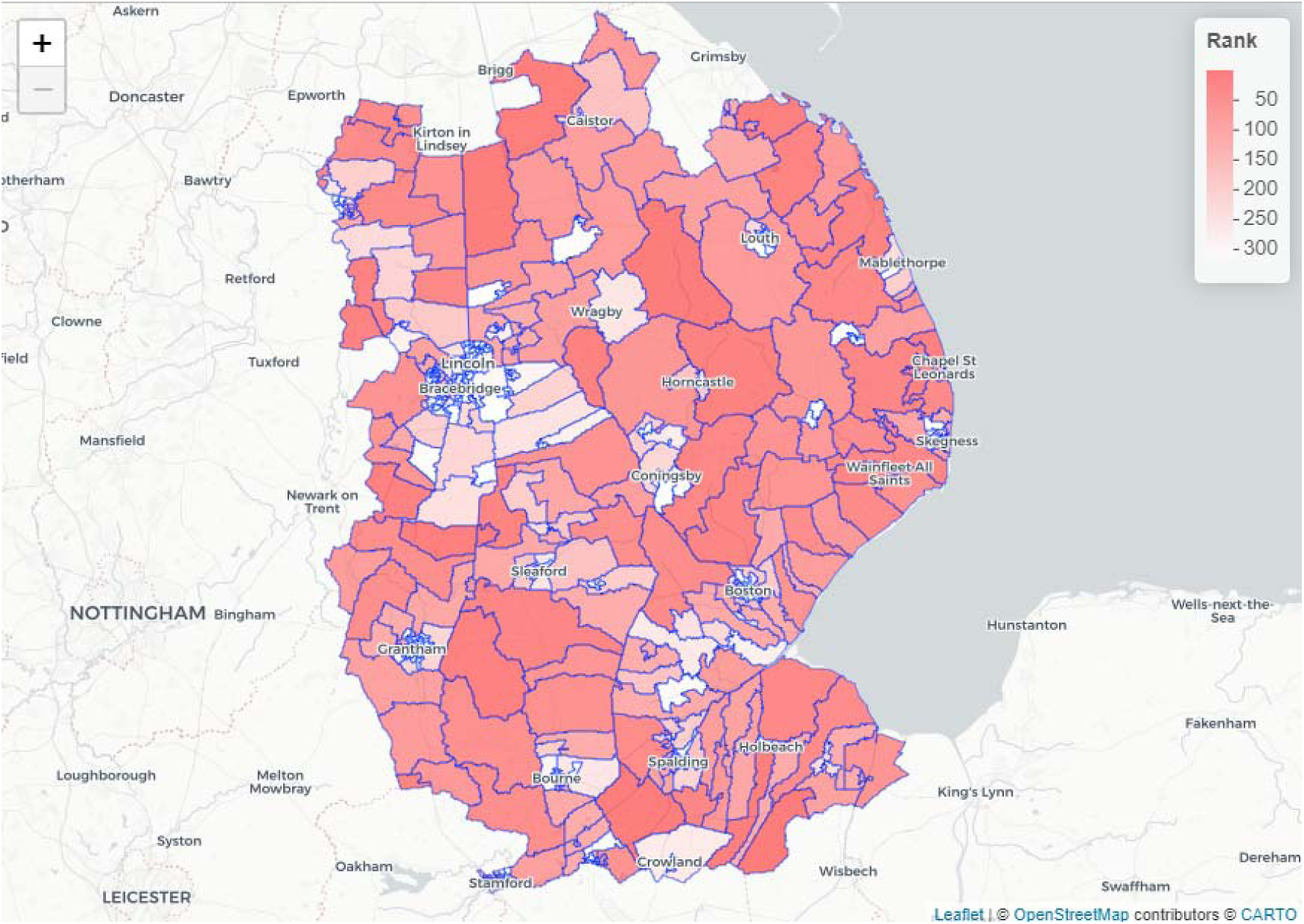
Map of Lincolnshire showing the relative rankings of the LSOA’s using factor analysis derived composite indicator scores for barriers to accessing GP practices and pharmacies (as defined by the travel times to access services by car or public transport) ranging from 1 (highest level of transport barrier – dark red) to 420 (highest level of transport barrier – white).

Figure 5 shows the combined effect of digital exclusion and transport barriers by dichotomising the LSOAs as high or low for each of the composite indicators. We see that in general the eastern coastal communities experience high digital exclusion and long journey times to access medical care. The exception to this being the major coastal towns, Skegness and Mablethorpe. A similar pattern is seen for a band of LSOAs running from Caistor in the north of the county towards Spalding and Holbeach in the South.

**Figure 5:**
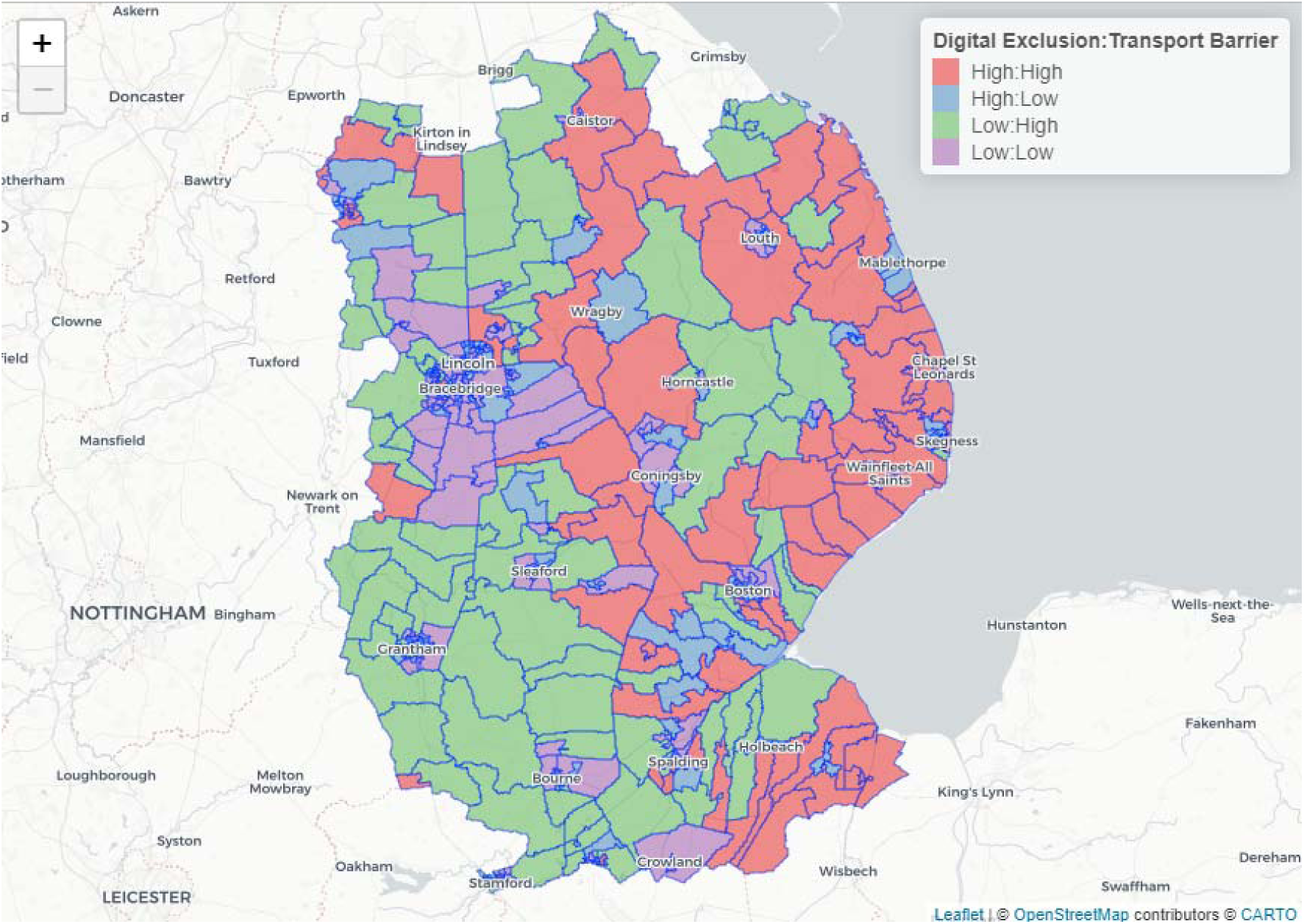
Map of Lincolnshire showing the combined effect of transport barriers and digital exclusion. The two rankings in Figures 3a and 3b were divided into high (above the mid-point) and low (below the mid-point) and all LSOA’s were then defined into four factorial groups for digital exclusion and transport barriers (high-high, high-low, low-high and low-low).

Within the overall trend of digital exclusion shown in Figure 3 there is a high degree of local heterogeneity. For example, within the city of Lincoln (Figure 6) or the town of Grantham (Figure 7), neighbouring LSOAs have very different levels of digital exclusion.

**Figure 6:**
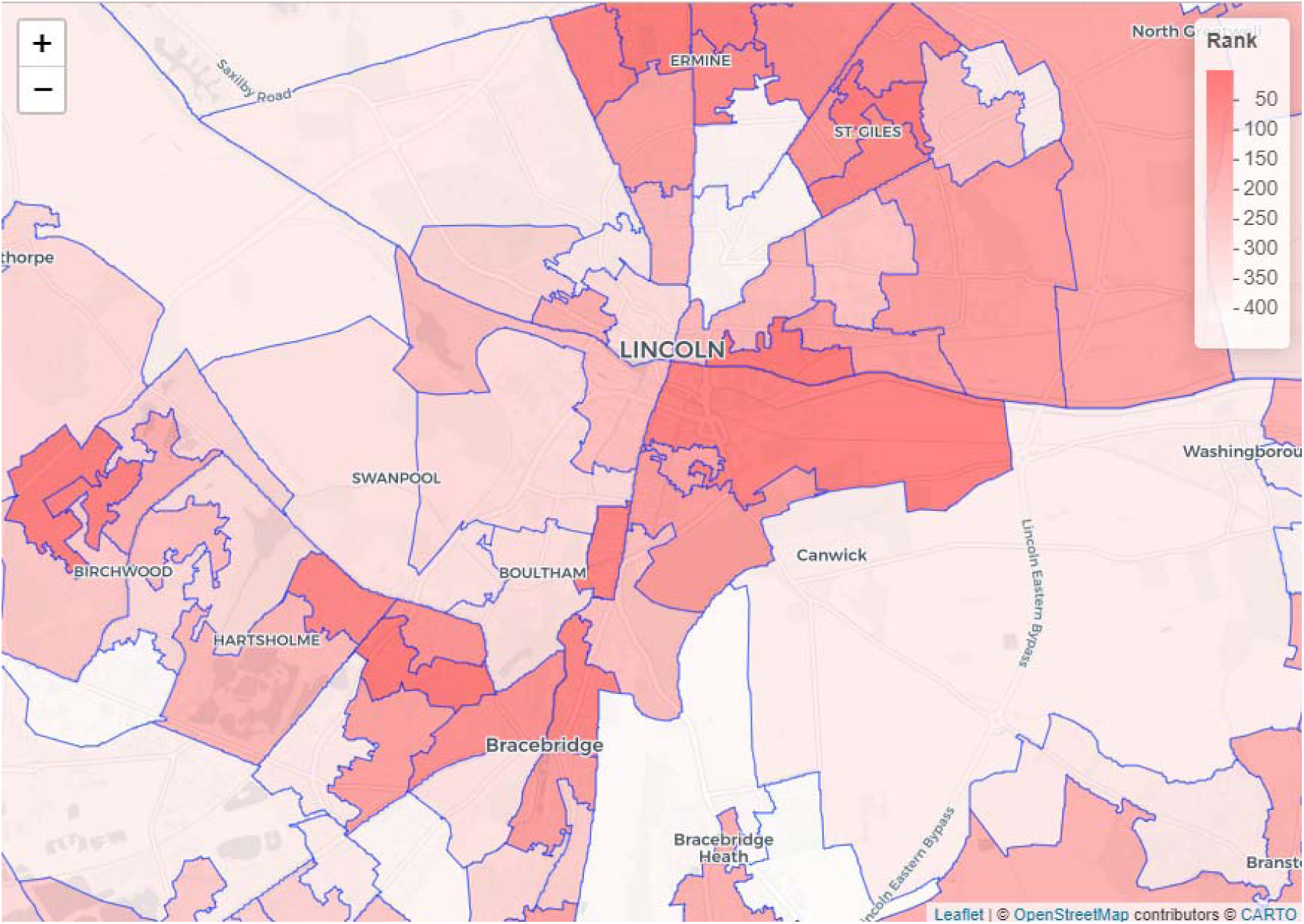
Expanded view of LSOAs in the city of Lincoln showing the local variation in the extent of digital exclusion.

**Figure 7:**
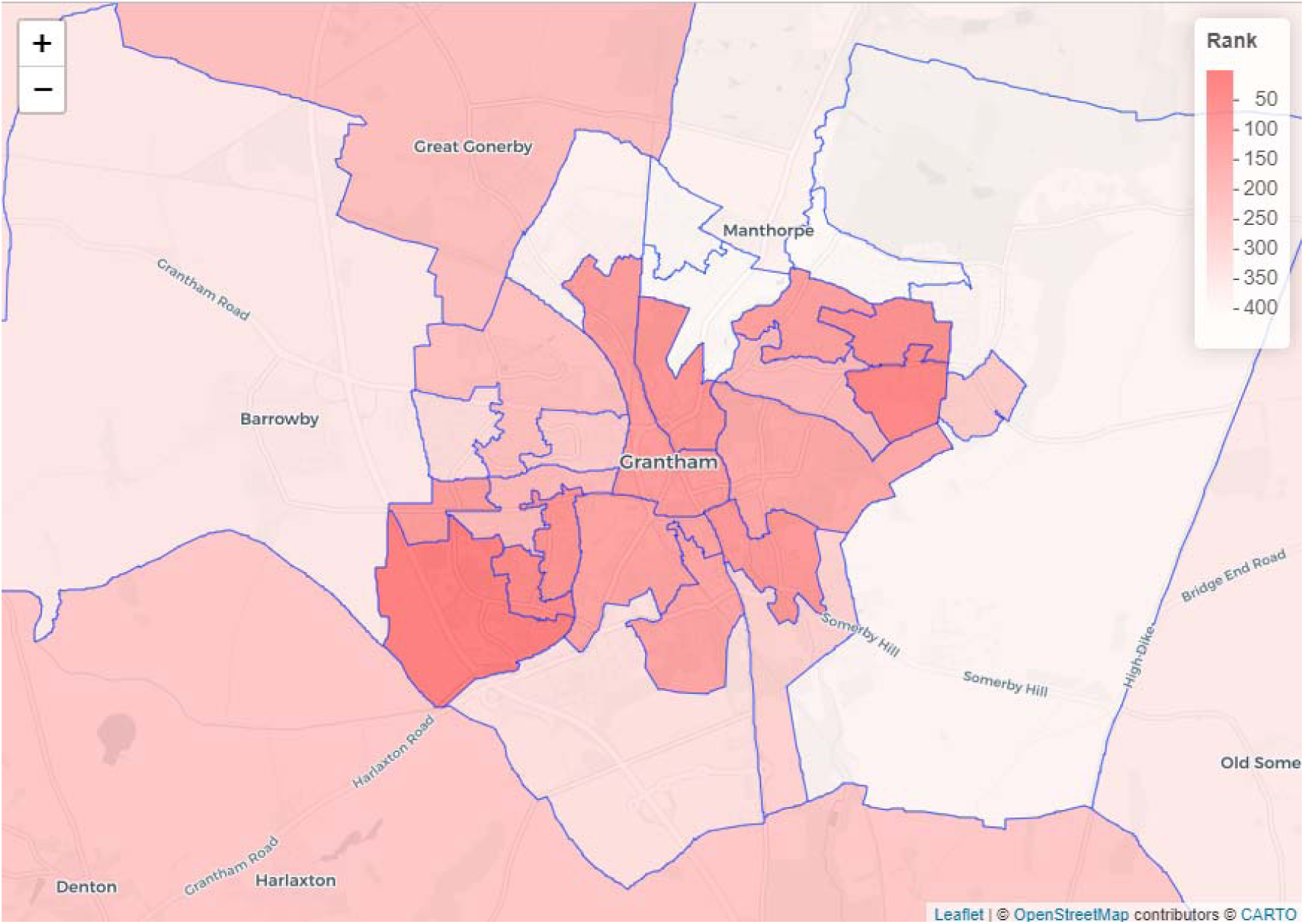
Expanded view of LSOAs in the town of Grantham showing the local variation in the extent of digital exclusion.

Iterative analyses indicated that it was optimal to extract three factors which explained over 63% percent of the variance in the original indicator variables for the Digital Exclusion composite (Table 3). Adding additional factors resulted in only a small increase in the explanatory power. The communality scores (Table 4) showed that these represented a reasonably high level of the variation in all but the INT (internet download speed) (communality = 0.102) and EXP (Composite of Experian Indicators) (communality = 0.207).

**Table 3:** Proportion of variance in the dataset explained by each factor.

|  | Factor 1 | Factor 2 | Factor 3 |
| --- | --- | --- | --- |
| Proportion of variance explained | 0.382 | 0.141 | 0.110 |
| Cumulative proportion of variance explained | 0.382 | 0.523 | 0.633 |

**Table 4:** Factor analysis – Communality and Factor Loadings (codes are as described in Table 1).

| Indicator | Communality<sup>9</sup> | Factor Loadings<sup>10</sup> |  |  |
| --- | --- | --- | --- | --- |
|  |  | Factor 1 | Factor 2 | Factor 3 |
| NQR | 0.571 | 0.300 | 0.363 | 0.591 |
| GPC | 0.684 | 0.820 | 0.107 |  |
| UNP | 0.879 | 0.912 |  | -0.199 |
| DER | 0.995 | 0.930 |  | 0.351 |
| LAC | 0.995 | 0.297 | 0.856 | 0.417 |
| INT | 0.102 | -0.146 |  | 0.269 |
| EXP | 0.207 | -0.323 | 0.300 | 0.116 |

An analysis of the individual factor loadings (Table 4) would suggest that the first factor is predominantly constructed from three indicators which show a high degree of mutual correlation (GPC, UNP & DER – loadings = 0.820, 0.912 and 0.930 respectively), with LAC, EXP and NQR making relatively lower contributions (loadings = 0.297, -0.323 and 0.300 respectively). This factor explained 38.2% of the variance in the original data (Table 3). Factor 2 explained a further 14.1% of the variance of the data. The dominant contribution to Factor 2 came from LAC (loading = 0.856) with a lesser contribution from NQR, GPC and EXP (loadings = 0.363, 0.107 and 0.300 respectively). The highest loadings for Factor 3 came from NQR (0.591), LAC (0.417) and DER (0.351), this factor explained a further 11% of the variance in the data.

### 4.1 Online Dashboard

An online dashboard the ‘Lincolnshire Digital Health Toolkit’ (https://lhih.org.uk/lincolnshire-digital-health-toolkit/) has been developed which presents data on the individual indicators and the composite metric superimposed on an interactive map (Figure 8). This enables users to interrogate factors associated with digital exclusion at an LSOA level alongside additional contextual information. This is designed to provide an analytical platform for local decision makers informing strategic approaches in the development of targeted interventions.

**Figure 8:**
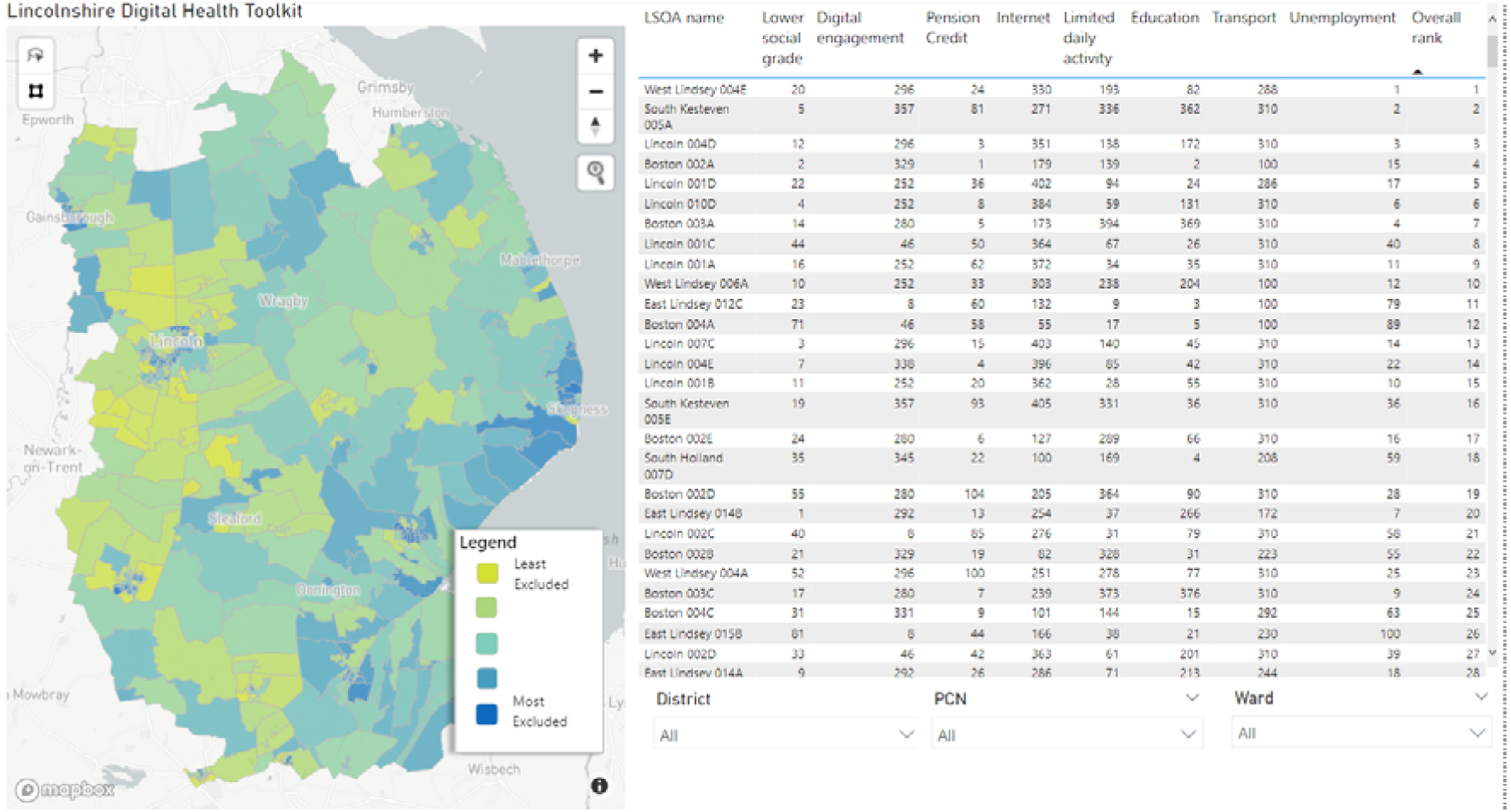
A screenshot from the Lincolnshire Digital Health Toolkit showing the LSOAs across the county categorised into 5 quintiles of digital exclusion based on their ranks in the composite indicator.

### 4.2 Case Studies

#### Targeted provision of digital devices

The Lincolnshire Digital Health Toolkit has been utilised by Lincolnshire County Council to fund a local charity in their delivery of services aimed at supporting those who are at a greater risk of digital exclusion. The index assisted with targeting the limited support and prioritising how this support is delivered. This included the provision of devices, data SIM cards, and help to access the internet to over 100 people in the community.

#### Provision of digital access kiosks in healthcare centres

It has been identified that whilst many people using medical practices access websites and social media via their mobile phones, they do not make use of digital services to support their healthcare provision. Anecdotal evidence from medical practitioners indicated that many of their appointments could be managed by people having access to digital services and information which is available across the health and social care sector. Using data from the Lincolnshire Digital Health Toolkit. digital access kiosks were placed in NHS properties located in or close to the most digitally deprived LSOA’s in the county.

## 5. Discussion and Conclusions

The rapid switch to online provision of services and interpersonal communication necessitated by the COVID-19 pandemic increased the level of digital exclusion experienced by some sub-groups such as those with low levels of digital literacy or access to appropriate digital devices [3]. The impact of digital exclusion in these groups must be addressed through both digital education initiatives and the provision of access to digital devices to reduce marginalisation and isolation. Metrics to quantify relative local differences in levels of digital exclusion are a necessary pre-requisite for the targeted provision of interventions to address this. The Lincolnshire Digital Health Toolkit provides a novel composite metric tailored to the conditions of this largely rural county and an interactive dashboard to support decisions on resource allocation.

There is a link between social deprivation and digital exclusion [44], however it is clear that other factors related to poor digital access or lack of IT skills are also important over and above the background level of deprivation in a particular community. In this study, methodologies for the development of indicators of intra-country economic performance and social development have been used to develop a composite index of digital exclusion which balances the different underlying constructs that drive exclusion.

We have identified three underlying constructs in the individual indicator variables that were combined to create this composite. The first and dominant factor can be described as representing socioeconomic deprivation and is composed of the highly correlated indicators describing the proportion of those in each LSOA: claiming guaranteed pension credit, unemployed and with lower socioeconomic status. The predominant factor in the second construct is lack of activity with a smaller contribution from the Experian score and having low levels of qualifications. The largest contribution to the third construct is again low levels of qualifications, as the majority of other indicators also contribute with somewhat lower loadings it is more difficult to define what concept this represents.

Increasingly, alternative access to primary care in the UK is available via either online consultation or face to face appointments. It may be argued that those who live close to a medical facility will, to some extent, have the means to overcome digital exclusion. Those living in rural communities on the coast and in the central region of Lincolnshire were found to experience the combined impact of digital exclusion and of having to travel significant distances in order to access medical facilities. This combined analysis may be particularly useful in targeting interventions to reduce digital exclusion to those areas of greatest need.

Digital exclusion is a multi-faceted concept. Individuals may experience exclusion from certain services but not from others for a variety of reasons [45]. For example, the high prevalence of smartphone ownership, at least among those in younger age groups, may mean that barriers to accessing social media and instant messaging applications are relatively low. This same group may, however, experience barriers when accessing more complex systems such as those used to apply for employment or access government and council services, due to a lack of IT skills, equipment, or adequate broadband. It is important to take a nuanced approach when interpreting a composite indicator for digital exclusion and explore local variations in the individual indicator variables from which the composite has been derived.

Aggregating the component indicators at the LSOA level may lead to a loss of ability to understand individual level experiences of digital exclusion. For example, in a single multi-generational household there may be younger members who are more internet aware, so called ‘digital natives’ living with those of an older generation who have had to adapt to digital technology later in life. Further development of this tool should include a multi-level algorithm for individual level and aggregate inputs to develop a more nuanced individual metrics of exclusion.

The use of the Experian indicators derived from representative data aggregated up to LSOA level did not add a significant amount of power to describe the overall variation in the underlying data and hence made only a small contribution to the composite. This may be due to the lack of discrimination between LSOAs in the underlying Experian indicators with many LSOAs showing identical values for the same indicator. However, the specific nature of the information given by these indicators may be useful in exploring fine-grained aspects of exclusion.

A fundamental question remaining is whether this or other metrics adequately reflect individual’s experiences of digital exclusion. In future studies we plan to carry out representative individual level surveys to validate and improve the predictive ability of the composite metric.

These indicators and the composite metric will allow those with responsibility for commissioning and planning the delivery of services, communicating, and engaging with local populations, and supporting people to access services to understand the likelihood of digital exclusion in an area and the factors which underly it. Services and activities could then be designed to lower barriers for those at higher risk of digital exclusion. The tool should be used alongside other local knowledge, expertise, and intelligence to support decision making.

## Data Availability

All code is freely available for download from the following GitHub repository https://github.com/Paul-Mee/digex_linc .Sources to access the underlying data are described in the manuscript.

## Footnotes

1 LSOAs are small areas designed to be of a similar population size, with an average of approximately 1,500 residents or 650 households. There are 32,844 LSOAs in England. They were produced by the Office for National Statistics for the reporting of small area statistics.

2 The individual indicators in the DERI score are; the proportion of the population in the oldest age groups (over 65s and over 75s) , the proportion of over 65s on pension credit , the proportion of adults with no qualifications, the unemployment rate, the Index of Multiple Deprivation which is itself a composite indicator describing various dimensions of deprivation, [46] the proportion of homes with slow broadband access (<30 and <10 MBits/s) and the average download speed.

3 The NRS social grades are a system of demographic classification used in the United Kingdom. They were originally developed by the National Readership Survey (NRS) to classify readers, but have become a standard for market research [35]. Level D in the classification is defined as ‘Semi-skilled and unskilled manual workers’ and Level E as ‘State pensioners, casual and lowest-grade workers, unemployed with state benefits only’. The other grades A, B, C1 and C2 are defined as higher grades of employment.

4 Pension Credit gives recipients aged over the state pension age and on a low income extra money to help with living costs. Source: Pension Credit: Overview - GOV.UK (www.gov.uk)

5 Connected Nations (Summer 2022 update): https://www.ofcom.org.uk/research-and-data/multi-sector-research/infrastructure-research/connected-nations-2022

6 Office of National Statistics (ONS)

7 Experian Mosaic is cross channel classification system used to segment the population based on a comprehensive list of different factors including demographics, behaviours, and employment for example. Lincolnshire County Council hold a license for Experian Mosaic.

8 gov.uk (https://www.nao.org.uk/wp-content/uploads/2020/10/Transport_accessibility_tool_Tech-guide.pdf)

9 Communality indicates the overall proportion of the variance in the indicator variable explained by the factors.

10 Factor Loadings are the contribution of each original variable to the factor.

